# Online attitudes toward mental illness anti-stigma efforts: A content analysis

**DOI:** 10.64898/2026.09.11.26362889

**Authors:** Anthony L. Colonello, Rosina Mete

**Affiliations:** School of Graduate and Postdoctoral Studies, Western University, London, Ontario, Canada; School of Health and Social Sciences, City University in Canada, Edmonton, Alberta, Canada; Faculty of Behavioural Sciences, Yorkville University, Fredericton, New Brunswick, Canada

**Keywords:** Social Stigma, Mental Health, mental illness, Reddit, social media, Social Media, Health Promotion

## Abstract

Reddit has emerged as an accessible, anonymous platform for public discourse surrounding mental illness, yet little research has examined how users perceive and evaluate existing anti-stigma initiatives. This study addresses this gap by analyzing public discourse around destigmatization efforts, including both corporate-led campaigns (i.e., Bell Let’s Talk) and broader community-based discussions. We conducted an interpretive content analysis of Reddit posts and comments identified through an a priori search protocol registered on the Only Science Framework. We drew a criterion-based purposive sample from r/mentalhealth and general Reddit using keywords related to anti-stigma discourse (e.g., “anti-stigma,” “destigmatizing,” and #BellLetsTalk). Fifty posts met inclusion criteria, yielding 581 coded references. Posts predominantly addressed mental health in general terms rather than specific diagnoses, with public stigma (n=28 posts) and structural stigma (n=16 posts) most discussed; internalized stigma was rarely a primary focus. #BellLetsTalk posts located outside the mental health community generated substantially greater engagement than posts within r/mentalhealth, with several threads exceeding 300 comments. Four themes emerged during the qualitative analysis: (1) anti-stigma efforts are unhelpful (n=381), (2) hopeful sentiments about anti-stigma efforts, (3) the mixed role of media, and (4) future directions for anti-stigma efforts.

## Introduction

Social media platforms are often used to disseminate mental-health-related content and to facilitate discussions around mental illness (1–3). Interdisciplinary literature has widely studied a variety of digital platforms, including the popular online discussion forum Reddit (4). There is a range of research examining phenomena such as how mental illness is constructed online (5), how social media influences mental health (6), and how health information is disseminated online (7). The unique features of Reddit, such as accessibility and anonymity make the online community an ideal venue to have conversations about vulnerable or stigmatized topics, such as mental illness (8).

These digital spaces offer people the opportunity to access both support and perspectives regarding psychological concerns and symptoms. Regarding stigma towards mental illness, mass media anti-stigma campaigns have a small to moderate effectiveness, which is limited by a lack of long-term follow-up (9). There is a dearth of research examining the public discourse surrounding the perceived usefulness of and attitudes towards current efforts to reduce the stigma against people with mental illness. This study aims to address the gap by analyzing a sample of the current discourse surrounding destigmatizing mental illness. We will present a mixed-methods analysis.

### An overview of anti-stigma mental illness campaigns

Over the past two decades, public awareness campaigns addressing mental health stigma have gained considerable momentum in North America. In Canada, Bell Let’s Talk stands out as a prominent corporate-led initiative, launched in 2011 by Bell Canada Enterprise Inc. The campaign includes an annual awareness day in January, during which Bell donates five cents to mental health programs for every eligible text message, call, tweet, or other social media interaction that uses the campaign hashtag #BellLetsTalk. Since its inception, Bell Let’s Talk has raised hundreds of millions of dollars for Canadian mental health initiatives and has stimulated extensive public discussion about depression, anxiety, and related conditions (10). Nevertheless, scholars have expressed concerns that corporate-sponsored campaigns may contribute to the commodification of mental health advocacy and have questioned whether fundraising metrics effectively translate into reduced stigma at the community level (11).

In the United States, large-scale anti-stigma initiatives have been implemented by organizations such as the National Alliance on Mental Illness (NAMI) and the Substance Abuse and Mental Health Services Administration (SAMHSA). Campaigns such as “StigmaFree” seek to normalize help-seeking behaviour and challenge harmful stereotypes. These initiatives employ a contact-based education model, which emphasizes the lived experiences of individuals with mental illness as a primary means of shifting public attitudes. Research consistently demonstrates that direct or indirect contact with individuals who have personal experience of mental illness is among the most effective strategies for reducing both social and self-stigma (12,13). Public health messaging following this approach has been adapted for television, print, and digital platforms to reach diverse demographic groups nationwide.

Television and digital commercials have significantly influenced public attitudes toward mental illness by providing accessible entry points for audiences who might not otherwise engage with mental health discourse. Campaigns such as the Ad Council’s partnership with NAMI have produced widely circulated public service announcements that portray individuals in recovery and encouraged viewers to start conversations with loved ones and seek professional support. In Canada, organizations such as the Centre for Addiction and Mental Health (CAMH) have also created emotionally resonant media content intended to humanize psychiatric conditions and challenge associations between mental illness and dangerousness or incompetence (14). Communication scholars have analyzed how the framing of these commercials, especially the focus on hope, recovery, and social connection, can influence audience perceptions (15). However, critics argue that brief media exposure alone is rarely sufficient to produce lasting attitudinal change without concurrent structural and policy interventions.

### Reddit and mental health discourse

Online communities, particularly Reddit forums, have emerged as informal yet influential spaces for reducing mental health stigma and providing peer support. Subreddits such as r/mentalhealth, r/depression, and r/anxiety foster environments where users share personal experiences, offer mutual support, and challenge stigmatizing narratives in real time. In contrast to institutional campaigns, these forums are primarily user-generated and facilitate a degree of authenticity and vulnerability that broadcast media rarely achieves (16). While concerns about misinformation, unmoderated content, and lack of clinical oversight are valid, the reach and accessibility of these forums suggest that they play a meaningful, though undertheorized, role within the broader landscape of anti-stigma communication.

## Methods and materials

We will conduct an interpretative content analysis (17) of a sample of posts and comments identified from an *a priori* search protocol. The search will be registered on the Open Science Framework prior to data collection. An interpretative content analysis includes both qualitative and quantitative components. It encompasses both the manifest and latent content that exists within a text. Interpretive content analysis extends beyond traditional descriptive content analysis by offering deeper perspectives in answering more inferential questions. For example, it aims to answer questions like “to what effect” and “for whom” (p. 59). Consistent with the content analysis, we will present a quantitative extraction of key characteristics from the selected data, including year of post, author background (i.e., lived experience, mental health professional, or general public), the types of stigma discussed in posts (i.e., structural, internalized, or public stigma) and the number of upvotes on posts.

### Data collection

Cross-sectional searches of Reddit (https://www.reddit.com/) were conducted between June 27^th^ 2026, and August 22^nd^ 2026, using the website’s native search feature. We have developed an *a priori* inclusion and exclusion protocol to guide our criterion-based purposive sampling. Norman Adams (18) highlights that data can be collected from Reddit using both automated scraping methods (e.g., APIs, Pushshift) or manual search-and-screen approaches, depending on the aims of a study’s methodology. Given the qualitative nature of our study, conducted a manual search using the search procedures model after two studies by Slemon and colleagues (19,20). This included developing a protocol which first identifies subreddits to search, establishes search terms, and identifies inclusion and exclusion criteria. We searched each keyword against the inclusion and exclusion criteria outlined in Table 1.

**Table 1.** Inclusion and Exclusion Criteria.

| Inclusion Criteria | Exclusion Criteria |
| --- | --- |
| 1. Posts that mention attitudes about the nature of stigma towards people with mental illness<br>2. Text containing a minimum of 100 words collectively (including posts and discussions)<br>3. Reference to any psychological disorder or symptom (e.g., depression, bipolar, schizophrenia)<br>4. Posts pertaining to both adult and youth mental illness<br>5. All geographic locations<br>6. Posts in English<br>7. Posts within the last 8 years | 1. Posts not involving or describing mental illness anti-stigma efforts<br>2. Posts that do not inquire about opinions or attitudes about anti-stigma efforts<br>3. Text containing less than 100 words collectively (including posts and discussions)<br>4. Duplicate or cross posted posts<br>5. Posts in languages other than English<br>6. Posts or comments only including links to external sources (e.g., blogs, fundraisers) |
| **Note.** Any identifying information will be de-identified upon extraction and analysis. References to specific institutions will be anonymized when possible. |  |

Consistent with other research(21), we searched keywords on the subreddit “r/mentalhealth,” which currently has approximately 310,000 weekly visitors. We manually copied the publicly available data to create a text corpus for analysis. Table 2 outlines the key terms searched for based on initial pilot searches of various terms related to the central research question. Additionally, we specifically searched #BellLetsTalk as it is a widely used anti-stigma initiative and social media hangtag (10). The dataset included only textual data, and other media forms (i.e., links and images) were excluded. The search filters were set to sort by “relevance” and “all time”, and it was searched on an account logged in to Reddit.

**Table 2.**
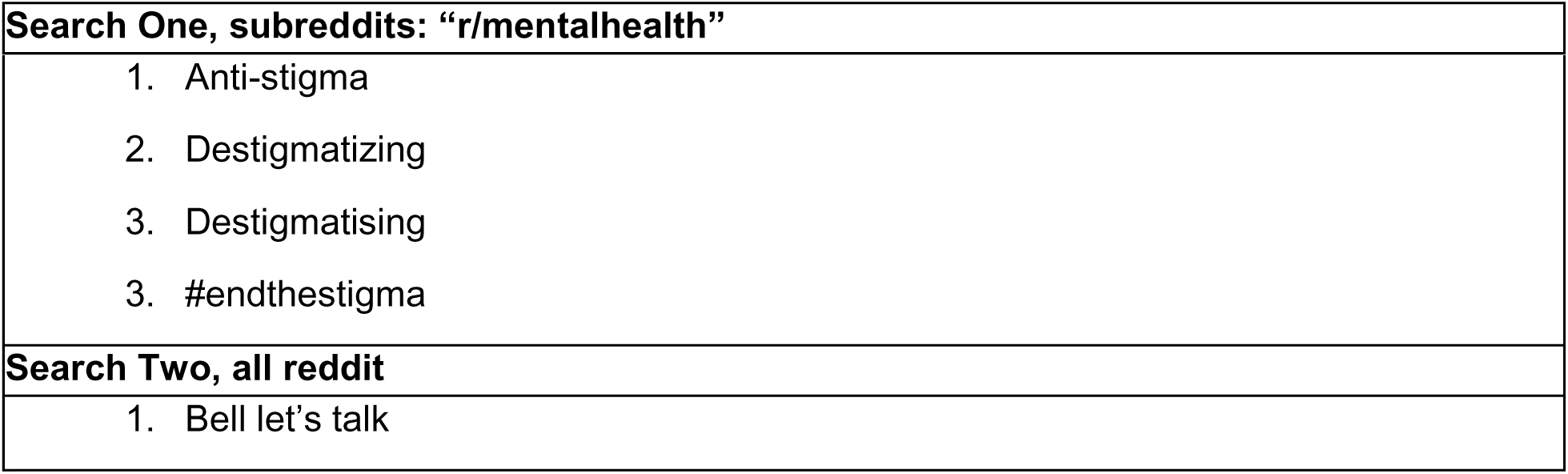
Key Search Terms.

### Data analysis

Two raters conducted the qualitative analysis and data extraction. We used inductive coding to identify key content and themes emerging within the data. The raters used both NVivo version 15 (22) and manual processes for coding and thematic development. The raters used an inductive coding approach with interpretive content analysis.

### Researcher Positionality

We sought to minimize the influence of our own views on the analysis by adhering to a structured coding framework and basing coding decisions on the data content. We recognize, however, that constructing any coding framework involves interpretive choices, and that our perspective is shaped by a clinical and mental health lens. Both authors are interdisciplinary mental health practitioners, educators, and researchers, with graduate training in counselling and psychology. This orientation informed how we conceptualized and organized the coding categories, but we did not treat it as data.

### Findings

Overall, we analyzed 50 posts to develop content and themes. We reviewed each post that met the search criteria, along with its comments. We extracted the manifest content to produce the characteristics of the data sample, and also conducted a qualitative interpretive analysis of latent content, which generated four findings. Table 3 outlines the posts included and specific information on diagnosis and stigma mentioned.

**Table 3.**
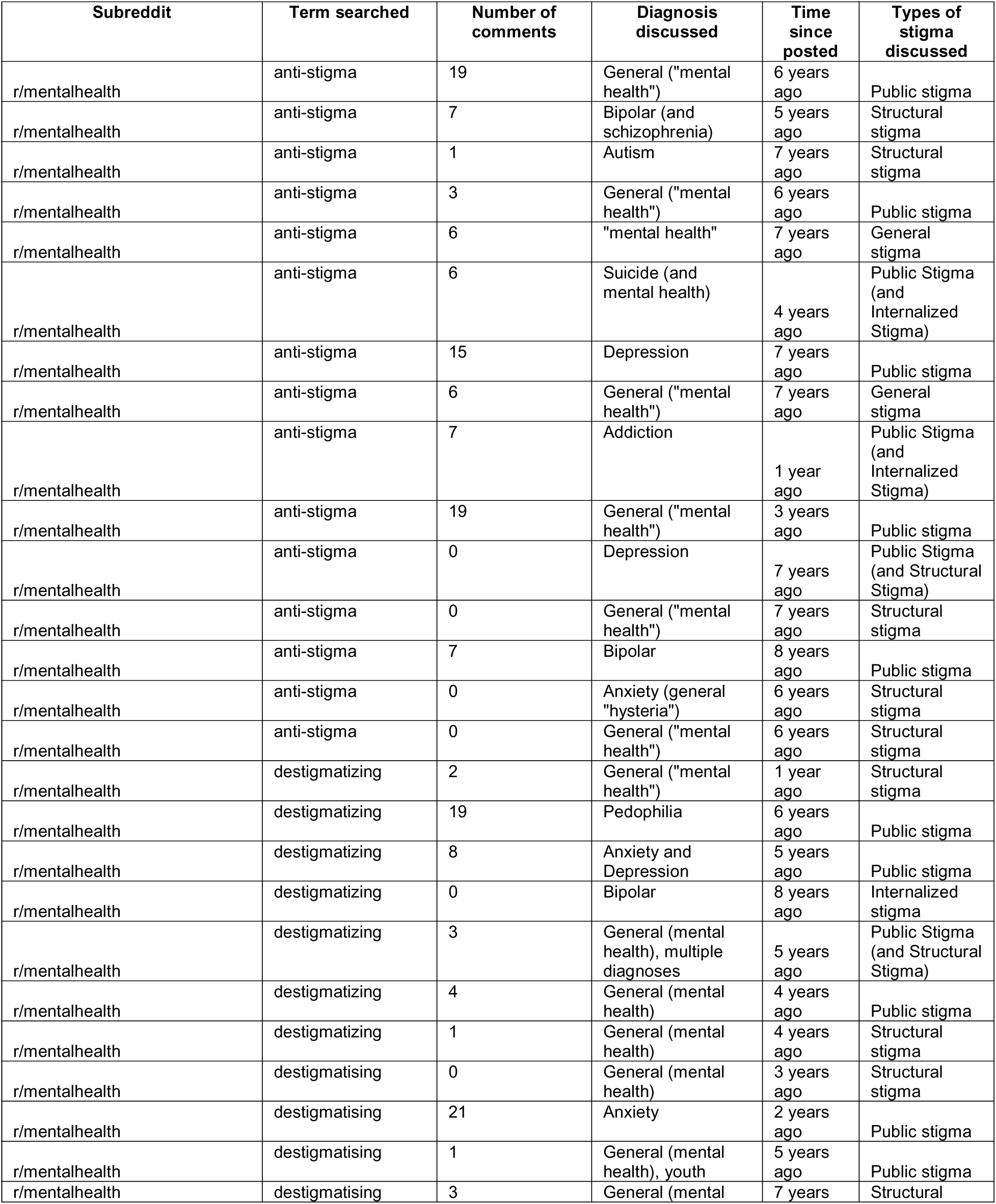
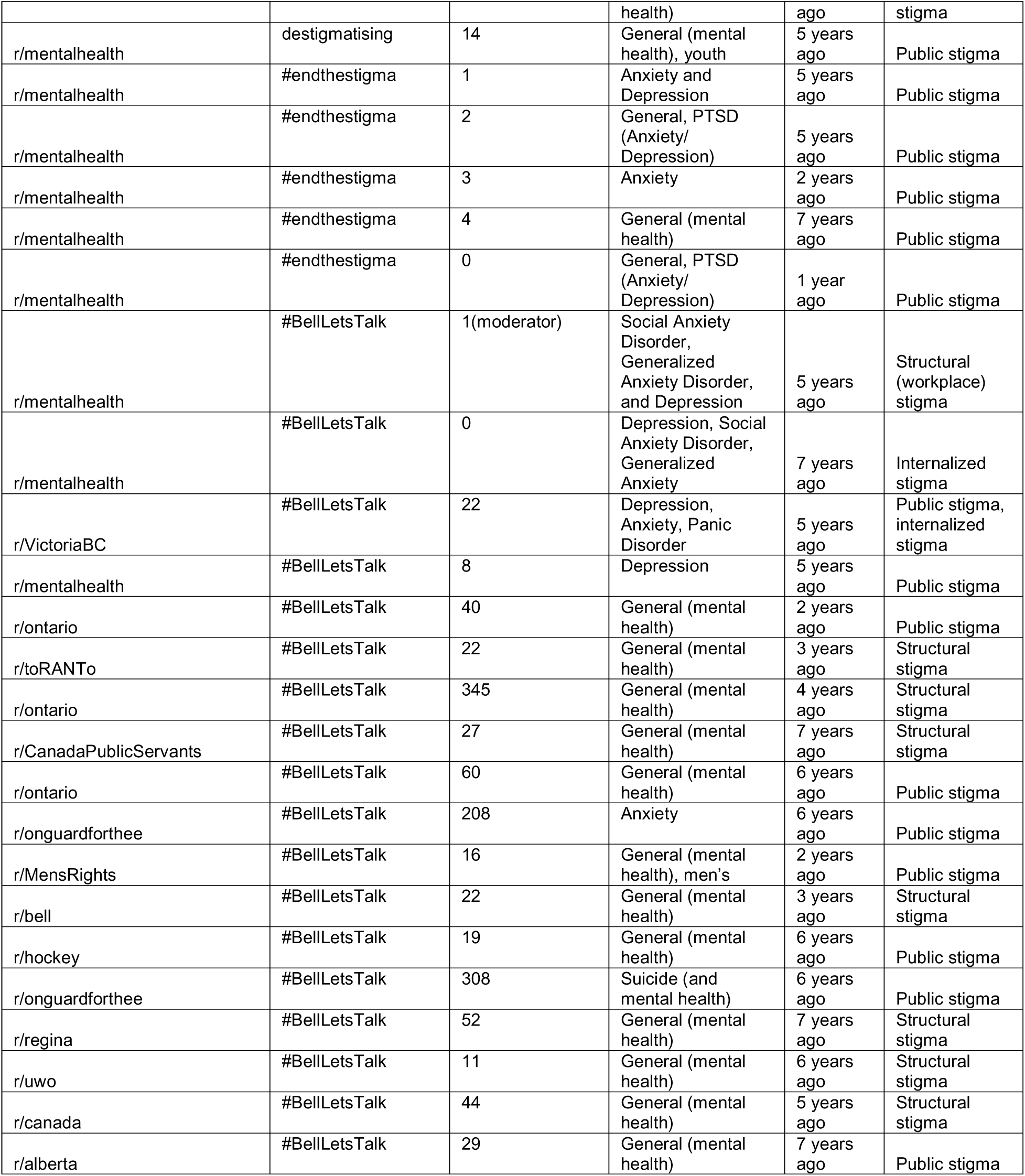
Expanded Characteristics of Included Search Results (n=50 posts)

The results demonstrated that #antistigma and #BellLetsTalk searches yielded the most posts within the designated timeframe. While the Bell Let’s Talk initiative first occurred well over a decade ago, the conversation has continued among Reddit users and in specific Canadian Reddit threads such as UWO (University of Western Ontario), Hockey, Regina, Alberta, and On Guard for Thee.

Overall, most posts were completed over four years ago with few in recent years. This may reflect that the Bell Let’s Talk discussion has become more mainstream within Canadian society. Posts were generally not recent. Most had been posted between four and eight years before data collection. Only seven posts were three years old or newer. Table 4 summarizes the combined characteristics of the included studies.

**Table 4.** Combined characteristics of included Reddit posts (n=50)

| Post Characteristics | n (%) |
| --- | --- |
| **Subreddit** |  |
| r/mentalhealth | 35 (70) |
| Misc. subreddits<sup>1</sup> | 9 (18) |
| r/ontario | 3 (6) |
| r/onguardforthee | 2 (4) |
| r/ bell | 1 (2) |
| **Search Terms Included** |  |
| "#BellLetsTalk" | 18 (36) |
| "anti-stigma" | 15 (30) |
| "destigmatizing" | 7 (14) |
| "destigmatising" | 5 (10) |
| "#endthestigma" | 5 (10) |
| **Timeliness (Years Since Initial Post)** |  |
| 1-2 years ago | 7 (14) |
| 3-4 years ago | 8 (16) |
| 5-6 years ago | 21 (42) |
| 7-8 years ago | 14 (28) |
| **Primary Diagnosis/ Condition Discussed** |  |
| General mental health | 27 (54) |
| Anxiety/ depression<sup>2</sup> | 12 (24) |
| Bipolar disorder | 3 (6) |
| Post-traumatic stress disorder | 2 (4) |
| Suicide/Suicidality | 2 (4) |
| Autism | 1 (2) |
| Substance use disorder | 1 (2) |
| Pedophilic disorder | 1(2) |
| Schizophrenia | 1(2) |
| **Type of Stigma Described in Main Post** |  |
| Public stigma | 29 (58) |
| Structural stigma | 17 (34) |
| Internalized stigma | 2 (4) |
| General stigma, unclear | 2(4) |
| **Initial Post Engagement Metrics** |  |
| Combined number of comments | 1,415 |
| Overall Net Vote Scores | 0 - 4,800<sup>3</sup> |
| <sup>1</sup>Misc. subreddits included: r/VictoriaBC, r/toRANTo, r/CanadaPublicServants, r/MensRights, r/hockey, r/regina, r/uwo, r/Canada, r/uAlberta.<br><sup>2</sup>Included a primary dialogue around anxiety, social anxiety, generalized anxiety, and panic disorders.<br><sup>3</sup>Based on a summed Net Vote Score of 14, 753 across all posts (n=50). Median = 5 and IQR = 28. |  |

### Diagnoses discussed

Most posts discussed mental health in general terms rather than naming a specific condition. Over half of the 50 posts (n=27, 54%) referred to “mental health” or “mental illness” broadly, sometimes alongside a passing reference to specific populations such as men or youth. Where specific diagnoses were named, depression and anxiety were the most common (n=12, 24%), followed by bipolar disorder (n= 3, 6%). Single posts addressed addiction, autism, suicide, and pedophilia. Posts tagged with #BellLetsTalk were especially likely to speak about mental health in general rather than about a particular diagnosis.

### Types of stigmas discussed

Public stigma was the most frequently discussed form of stigma, appearing as the primary focus in 29 posts (58%). Structural stigma was the primary focus of 17 posts (34%). Internalized stigma was the main focus of only two posts (4%), though it was mentioned as a secondary theme in three others. One post focused on workplace stigma, and two posts (4%) discussed stigma in terms too general to classify and were considered general stigma. Although the comments discussed a variety of forms of stigma (i.e., internalized, structural, and public), coding was limited to the primary type of stigma discussed in the main post. Authors categorized based on the essence of stigma discussion captured in the main post.

The type of stigma discussed varied by search term. Posts found through “#endthestigma” focused almost entirely on public stigma. Posts found through “anti-stigma” and “destigmatizing” were more evenly split between public and structural stigma. The clearest pattern appeared in the #BellLetsTalk posts. When these posts were located in r/mentalhealth, they tended to describe personal experiences of public, internalized, or workplace stigma. In general, Canadian subreddits, they more often addressed structural stigma, which in this context appeared to include commentary on the campaign itself and on Bell as a corporate sponsor.

### Engagement with posts

Most posts received few comments. In r/mentalhealth, the median post received fewer than ten comments, and nine posts received none. The #BellLetsTalk posts outside r/mentalhealth received far more engagement. Six of these posts received more than 40 comments, and the two largest threads, both in r/onguardforthee and r/ontario, received 308 and 345 comments respectively. In other words, the posts that generated the most discussion were not in the mental health community but in general audience communities responding to a national awareness campaign.

The data suggested that anti-stigma discussion on Reddit is concentrated on public stigma and on mental health in general rather than on specific diagnoses or on internalized stigma. Discussion of structural stigma was more common when a corporate campaign (Bell Let’s Talk) was the point of entry, and this discussion drew much larger audiences than posts originating within the mental health community.

### Qualitative analysis: thematic development

Four themes were developed from the data: 1) anti-stigma efforts are unhelpful; 2) hopeful sentiments about anti-stigma efforts; 3) the mix of media; and 4) future directions for anti-stigma efforts. For each theme, the coded reference number indicated unique experts from posters or commenters that represent the overall theme or subthemes. We coded 581 references across all themes.

#### Theme one: anti-stigma efforts are unhelpful

The most frequently occurring theme was that anti-stigma efforts are unhelpful (n=381 coded references). This included several subthemes: personal responses to the nature of stigma; preaching support without meaningful action; and stigma still remains. Comments coded under this theme questioned whether awareness campaigns change attitudes or improve access to care, and many treated corporate campaigns such as Bell Let’s Talk as marketing rather than meaningful action. Users were also critical of anti-stigma messages online, expressing frustration with awareness initiatives. This theme appeared most often in the large threads located in general Canadian subreddits, where discussion focused on structural problems such as wait times, funding, and workplace treatment that awareness alone does not address.

Some messages (n=25 coded references) specifically commented that public anti-stigma efforts are believed to be “virtue signalling,” which refers to actions that make people, groups, and/ or organizations appear virtuous. Virtue signalling and conceptual derivatives (i.e., “signal virtue” or “virtue signal”) appeared 55 times across two searches, including by the same users, some of which were coded together based on the discussion. Similarly, others in this theme (n=30 coded references) believed that anti-stigma campaigns promoted performative allyship. Users shared how, despite the messages about ending stigma, there remained structural barriers in the provision of mental health services and various types of stigma still impacted people’s daily lives.

#### Theme two: hopeful sentiments about anti-stigma efforts

A second, smaller theme (n=70 coded references) captured hopeful sentiments about anti-stigma efforts. These comments came mainly from r/mentalhealth and described awareness campaigns as a starting point for conversation, a way to feel less alone, or the reason someone first spoke openly about their diagnosis. Regarding Bell Let’s Talk, one user shared, “It does encourage people like us with mental illnesses to open. It’s better than absolutely nothing.” Several users similarly highlighted positive views about corporate initiatives, praising Bell Let’s Talk for the millions of dollars raised for mental health support and research. Others extended this hopeful sentiment to broader mental health destigmatization efforts, asserting that such public messages have “done leaps and bounds for how we receive care”. Messages in this theme also acknowledged that anti-stigma efforts are not perfect; however, they contrasted the current public mental health dialogue positively compared to previous generations.

#### Theme three: the mix of media

The third theme, the mix of media, described how users encountered anti-stigma messaging across several platforms at once. This theme included how media could support anti-stigma efforts (n=11 coded references) while also working against them (n=29 coded references). Media included both social and mainstream media (i.e., movies, news). Comments referred to television advertising, hashtags, celebrity involvement, and posts shared between social media sites, and users often evaluated the campaign based on where they saw it rather than on its content.

Critiques of media suggested that inaccurate portrayals of mental illness in shows, movies, or online can perpetuate stigma by reinforcing harmful stereotypes about mental illness. Users described how media may sensationalize or misrepresent the lived experiences and presentations of mental illness. Contrary to this, some users highlighted how when the depictions of mental illness are accurate and helpful, this reduced internalized stigma and public stigma based on reported experiences.

#### Theme four: future directions for anti-stigma efforts

The fourth theme, future directions for anti-stigma efforts (n=90 coded references), gathered suggestions for what should replace or improve current approaches. These included redirecting campaign spending toward services, addressing specific and less socially accepted diagnoses rather than mental health in general, involving people with lived experience in campaign design, and moving beyond public attitudes to address stigma in institutions. There were two main subthemes: 1) a need to normalize and 2) whose responsibility is it. Within the first, users described a noted gap in health care systems where stigma prevails. Some (n=8 coded references) expressed this by comparing physical health to mental health, asserting that the disparity in how mental health is often stigmatized needs to be addressed. Users offered few concrete solutions; however, some (n=15 coded references) reported a desire for diagnostic reform. These users felt that psychiatric diagnosis was inherently stigmatizing, especially when systematic barriers (i.e., conflicting diagnostic opinions or long waits for accessing diagnosis) exist. Across this theme, users supported reducing stigma while calling for approaches that go beyond awareness alone.

## Discussions

This study examined how Reddit users talk about efforts to reduce mental illness stigma. Across 49 posts, the most common theme was that anti-stigma efforts are unhelpful, and this theme was most visible in the largest and most active threads. Public stigma was discussed most often, followed by structural stigma. Internalized stigma was rarely discussed. Posts that discussed mental health diagnoses in further depth highlighted greater stigma. These findings add to a growing body of work that uses social media to study public attitudes toward mental illness, and they suggest that the online public views current campaigns with considerable doubt.

### Skepticism as the dominant response

The prominence of this unhelpful theme aligns with concerns already raised in the literature about corporate awareness campaigns. (11) questioned whether fundraising totals translate into lower stigma in communities, and users in this sample raised the same question more directly. Much of this criticism appeared in general Canadian subreddits rather than in r/mentalhealth, and it was often coded as structural stigma. Consequently, when users criticized Bell Let’s Talk, they tended to outline wait times, funding shortfalls, and workplace practices rather than highlight individual attitudes. This finding corresponds to the Lancet Commission on ending stigma and discrimination, which argued that awareness alone is insufficient and that structural change and social contact with people with lived experience are the interventions with the strongest evidence (Thornicroft et al., 2022). Users appear to have reached a similar conclusion on their own.

The comment counts reinforced this reading. Posts about Bell Let’s Talk in general audience communities received far more engagement than any post in r/mentalhealth. Saha et al. (2019) found that awareness campaigns produce large spikes in social media activity, but the content of that activity is not always aligned with the campaign’s goals. The present findings suggest that a campaign can succeed in generating conversation while the conversation itself turns toward criticism of the sponsor. This does not mean the campaign has no value. It does mean that discussion volume should not be treated as evidence of reduced stigma.

### Hope and the role of the mental health community

The hopeful theme was smaller but important. It appeared mainly in r/mentalhealth, where users described campaigns as a reason they first spoke about their own diagnosis or as a signal that they were not alone. This fits with evidence that mass media campaigns have small but real effects on knowledge and attitudes (9,23). It also fits with the view that online communities offer support that may not be available offline, particularly for people who fear disclosure (8,24). The difference between the two communities in this sample is worth noting. People who identify with a mental health community appeared to evaluate campaigns by their personal effect, while people in general communities evaluated them by their political and economic effect. Both evaluations are reasonable, and they are not in conflict. A campaign can help an individual feel less alone and still fail to change the systems that produce discrimination.

### The influence of online attitudes on destigmatization

Online attitudes matter for destigmatization in at least two ways. First, social media is now a primary source of mental health information for many people, and the tone of that discussion shapes what newcomers learn. Robinson and colleagues (25) found that mental health conditions were more often stigmatized and trivialized on Twitter than physical conditions. The present study found less overt stigma in these particular threads, which is not surprising given the search terms, but it did find widespread cynicism about anti-stigma work. This kind of cynicism may discourage participation in campaigns that do have modest benefits. Second, online discussion is a form of public feedback that campaign designers rarely receive through formal evaluation. The suggestions in the future directions theme, such as redirecting spending to services, involving people with lived experience, and addressing specific diagnoses rather than mental health in general, are close to the recommendations of current stigma research (26). The public and the research community appear to agree about what should change. The gap is between what is known and what campaigns actually do.

The focus on general mental health in most posts also deserves attention. Public attitudes toward depression and anxiety have improved over time, while attitudes toward conditions such as schizophrenia have improved little or have worsened (27). A campaign that speaks about mental health in broad terms may reinforce this pattern by drawing attention to the conditions that are already most accepted. Very few posts in this sample discussed psychosis, personality disorders, or substance use.

If online discourse mirrors campaign messaging, the conditions most in need of destigmatization may be the ones least discussed.

### Limitations and future recommendations

This study has several limitations. The sample was small and drawn from a limited set of search terms, so it does not represent all Reddit discussion about stigma. Additionally, data derived from Reddit users may not be generalizable to all populations, and the platform’s anonymity makes it impossible to verify who is engaging in the online dialogue. Accordingly, we also could not differentiate unique commenters, and the frequencies of thematic codes were relative to the overall extracted references.

Coding was interpretive, and other researchers could classify the same comments differently, or experts could possibly fit under multiple codes. To mitigate individual bias in this study, we held a series of meetings and discussions among the authors during theme development and after independent coding. Future research should compare discourse across platforms, track sentiment shifts across campaign years and examine how users with lived experience respond to campaigns differently from those without. Work that directly tests whether online criticism affects campaign participation or public attitudes would also be valuable.

Despite these limits, the findings suggest that the public is not opposed to reducing stigma but is doubtful that awareness campaigns in their current form can do it. This is also consistent with research recommendations (28) derived from a study exploring anti-stigma attitudes before and after a large-scale Canadian multimedia campaign, the 2009 Opening Minds campaign. The authors found that the over-1-million-dollar multimedia anti-stigma efforts (i.e., newspaper, television, social media) were ineffective at the population level, with no significant improvements in stigma knowledge, campaign awareness, or advocacy post-campaign. Similar to some recommendations captured in our review of Reddit (i.e., participants highlight how awareness is not enough, and expressed desire for organizational supports to address structural stigma in the workplace), Stuart and Pietrus (28)emphasize shifting anti-stigma efforts toward targeted interventions for schools, health providers, media, and workforces. However, this is one campaign and is not generalizable to all online public media efforts. Considering the discourse captured in our study, we argue that our findings reflect a desire from Reddit users for campaigns to extend beyond spreading a message or encouraging disclosure of mental illness. Campaign designers who take that doubt seriously may find in it a clearer sense of what the public expects from anti-stigma work.

## Data Availability

The study protocol was registered a priori on the Open Science Framework (OSF) prior to data collection. The full dataset generated and analyzed during this study is available from the corresponding author upon reasonable request.

## Acknowledgments

Authors have no conflicts of interest to report. The views represented in this data reflect online commentary on anti-stigma attitudes and campaigns and do not represent the personal views of the authors.

